# Young Maternal Age at first birth and Child Undernutrition in Bangladesh: Evidence from the Bangladesh Demographic Health Survey 2017-18

**DOI:** 10.1101/2023.08.27.23294684

**Authors:** Md. Alamgir Hossain, Md. Tariqujjaman, Novel Chandra Das, Rubaiya Matin Chandrima, S.M Hasibul Islam, Ahmed Ehsanur Rahman, Hassan Rushekh Mahmood, Aniqa Tasnim Hossain

## Abstract

**Background:** Childhood undernutrition is a serious public health issue that includes stunting, wasting, and underweight. In Bangladesh, many adolescent girls become mothers early. Giving birth at a young age is common, adversely affecting maternal health and child undernutrition. This study investigated the association between maternal age at first birth and undernutrition of under-five children in Bangladesh.

**Methods:** The study is cross-sectional and data was collected from BDHS 2017–18 survey. Mothers who were pregnant at the age of ≤ 19 years were regarded as young mothers and aged >19 years were regarded as adult mothers. Children were categorized as stunting (height for age), wasting (weight for height), and underweight (weight for age) based on the z-scores -2 as per WHO median growth guidelines. Multivariate logistic regression was employed to assess the association.

**Results:** This study revealed that 71.5% of mothers had their first child before 19 years old. The undernutrition status of the children of younger mothers and the children of adult mothers differed significantly, stunted (p<0.001), wasted (p<0.001), and underweight (p<0.001)).

Children of mothers aged under 19 years at their 1st childbirth were 1.4 times more likely to be stunted [Adjusted odds ratio (AOR): 1.4, 95% CI: 1.02-2.46; p=0.008], 1.6 times more likely to be wasted (AOR: 1.6; 95% CI: 1.09-1.78; p=0.039), 1.5 times more likely to be underweight (AOR: 1.5, 95% CI: 1.15-2.07; p=0.004) compared to children of mothers who are adults.

**Conclusion:** This research will draw attention to the policymakers taking the necessary initiatives focus on the maternal age at the time of first birth, which will help reduce all forms of undernutrition in under-five children.

## Background

Undernutrition is the term that refers to a person’s inability to meet their needs to maintain good health (1). Adequate nutrition is helpful for growth and progress but malnutrition threatens human life (2). Globally, in 2020, 45 million under-five children were wasted and 150 million were stunted (3) and 47 million were underweight (4). In low and middle-income countries (LMICs), childhood undernutrition which includes stunting, wasting, and underweight, is a serious public health issue. In LMICs, undernutrition is a factor in about 45% of under-five fatalities. (3). In South Asia; the prevalence of undernutrition is 15.7% (5); 8.2% were wasted, and 27.4% were stunted (6). Bangladesh made some progress on the prevalence of stunting (31%) (7), but this is still higher than the average (21.8%) for the Asian region (8). In Bangladesh, the prevalence of wasting (9.8%), but this is still higher than the average for the Asian region (8.9%) (8), and 21.9% are underweight in Bangladesh (7, 8).

Globally, around 1.8 million children die each year due to stunting and wasting, and 12% of the years are adjusted for disability (9). Undernutrition in childhood is a factor that affected cognitive development and causes brain damage, harming children’s motor development and investigative behavior (10). Undernutrition in children has been associated with atypical behavior, low academic performance, and mental retardation (11).

Young mothers are at risk of giving birth to undernourished children (12). The age at first birth refers to the percentage of mothers giving birth at certain ages. In this context, “young mother” refers to becoming a mother between the ages of 10-19 years (13). Women aged 15-19 who have given birth their first child are taken into account as per BDHS-17/18 (7). In LMICs, it is anticipated that per year 21 million of mothers aged between fifteen to nineteen become pregnant, and 12 million give birth (14). In South Asia, about one in two adolescent girls married before the age of eighteen and one in five girls give delivery before their eighteen years (15). As per the world bank, 37% of girls married before their eighteen years, and that impacted child mortality and nutrition (16). In Bangladesh, sixty-six percent of women gave birth before age twenty (7). Young mothers often suffer from abortion, ectopic pregnancy, miscarriage, maternal infections and obstructed labor (17). Birth outcomes of adolescent mothers are born with stillbirth, low birthweight, preterm birth, child mortality, severe neonatal conditions, birth injuries, and retarded growth (18-21).

In developing countries, young pregnancies mostly happened in economic insolvency, with less education, and among the rural population (22, 23). Approximately 71% of new mothers start breastfeeding, but due to labor and delivery problems, and low social support breastfeeding initiation was hampered (24). Adolescent mothers are less likely to get immunized, obtain regular health care, overfeed and underfeed their infants, and introduce solid early and inadequate nursing to their offspring (18, 25). Deaths of live newborns are fifty percent higher among babies of young mothers compared to mothers aged 20-29 years (22). The young mother requires special attention as she is at risk of adverse health outcomes rather than the older (26). A prospective study found that young mothers’ age (≤ 19 years) was associated with infants’ low birth weight, preterm birth, stunting, and failure of completing secondary school to the child (27). In developing countries, maternal and child undernutrition is caused by several factors, including a lack of suitable feeding habits for babies and young children (28). Even though the government has taken steps to discourage young marriage, it is yet common in rural and underprivileged areas. Early marriage influences becoming a mother at a premature age.

However, as per our knowledge no studies explored the association between young maternal age at their first birth and undernutrition in children in Bangladesh. We aimed to investigate maternal age at first birth and child undernutrition in Bangladesh using nationally representative cross-sectional health surveys (BDHS-17/18) data.

## Methodology

### Data source

Data extracted from the Bangladesh Demographic and Health Survey (BDHS)-2017/18. The two-stage stratified sample design used in the BDHS survey across all administrative divisions. Every division is further subdivided into zilas and every zila into upazilas. Subdivisions of the upazila serve as the main sample units (enumeration areas, EAs), wards and mohallas in urban areas, unions and mouzas in rural areas. EAs were chosen in the initial step with probability based on size. A sampling frame for the second step of household selection was created by completing a household listing operation in each selected EAs. An average of 30 households per EA were chosen in a systematic random sample during the second sampling step. The BDHS provides essential mother and child health, and nutrition information that represented at the national and divisional levels (7, 29).

### Samples

Child anthropometric measurements were the key outcome factors. Enumerators measured weight, height, and length using the internationally accepted standard protocol. Each child has measured height for age, weight for height, and weight for age to calculate Z-scores for comparison to the age and gender-appropriate child growth guidelines of WHO (30).

After requesting permission from the DHS program, datasets were downloaded from the DHS website. After excluding missing data, height limits and flagged cases (inconsistent, irrelevant and not applicable response etc), absence for height and weight measurement, and deaths, finally 7,643 children were included in this study (Fig. 1). Detailed information on the survey (such as the sampling method, the determination of the sample size, and the data collection procedure) is described in the BDHS 2017-18 report (7).

**Fig. 1.**
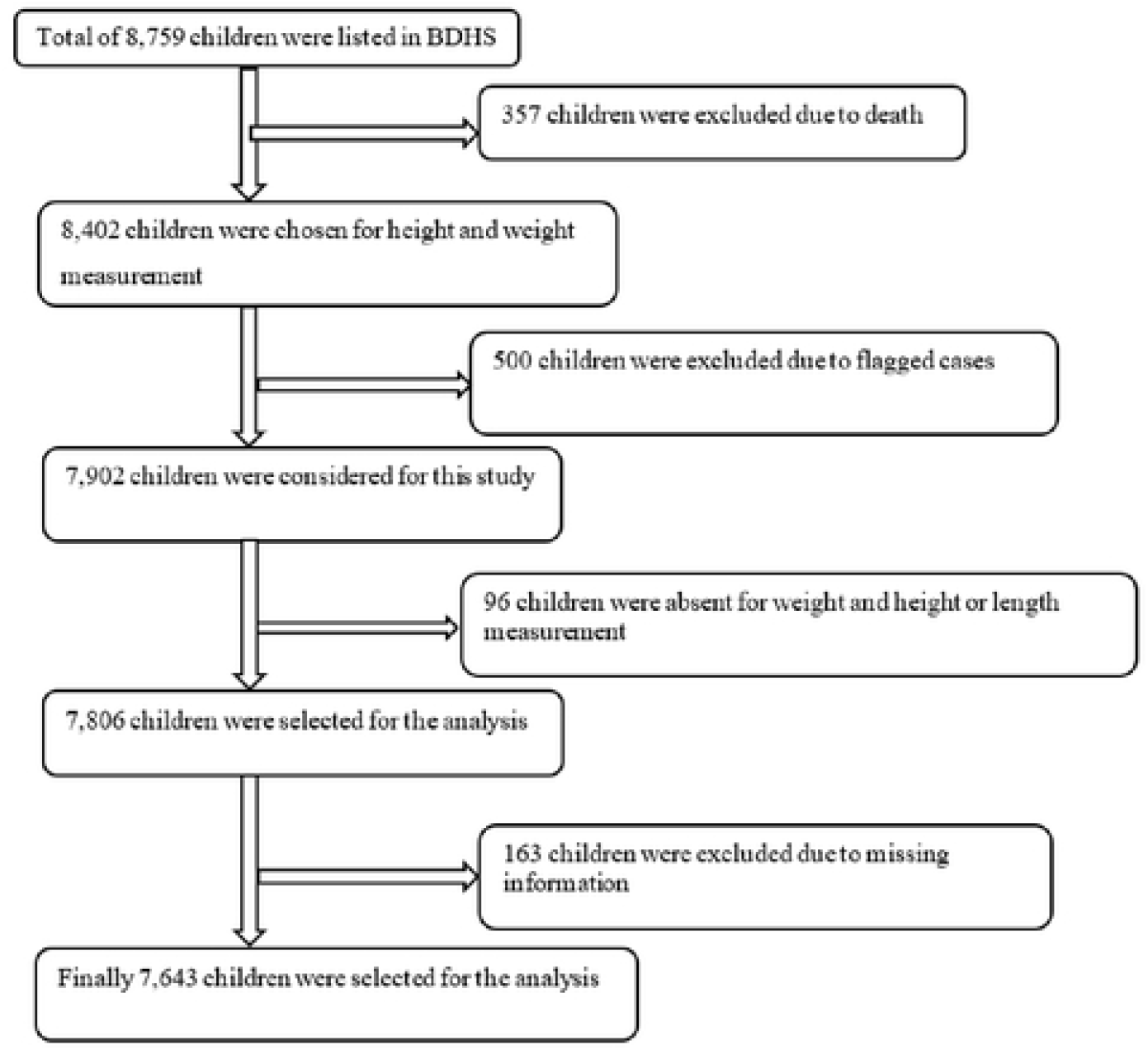
Sampling flow chart for the selection of the participants.

This study considers the maternal age at first birth as the primary exposure variable. We divided the ages of the mothers at their first birth into ≤ 19 and >19 years. To evaluate the link between maternal age at the time of the first birth and undernutrition in children. In addition, we took into account the mothers’ age (≤ 19 years, and >19 years), the household head (mother, husband, father, fathers-in-law, and others), the mothers’ body mass index (BMI) [underweight, normal weight, and overweight and obese (BMI 25->29.9 kg/m2)], the age group of the children (≤ 24 months, >24 months), the delivery by cesarean section (yes, no), the mothers’ education ((yes, no).

### Statistical analysis

The statistical program Stata (version 15) was used to analyze the data. Descriptive statistics were calculated and reported in percentages and weighted percentages. The bivariate analysis used the chi-square test to assess the significant associated factors of childhood undernutrition. Finally, after adjusting potential confounders, the association between undernutrition in under-five children and the mother’s age at first birth was examined using multiple logistic regression analysis.

### Ethical consideration

DHS data are publicly available upon justifiable request and are in the public domain. We did not need further ethical approval because the study protocol was accepted by both the Bangladeshi Ethics Commission and ICF International (7).

## Results

### Socio-demographic characteristics

About 72% of mothers’ age at first birth was below 19 years, 56.3% of household head was husband, 60.5% of women were of normal weight. About 50% of mothers completed secondary education, whereas 33.3% of fathers completed primary education. In the wealth index, we found that the distributions of poorer (20.4%) and richer (20.3%) and rest are almost similar. About 35% of mothers delivered babies by C-section. 15.7% of children were under 2500g at birth, and 40.2% of mothers were employed at the time of data collection (Table 1).

**Table 1:**
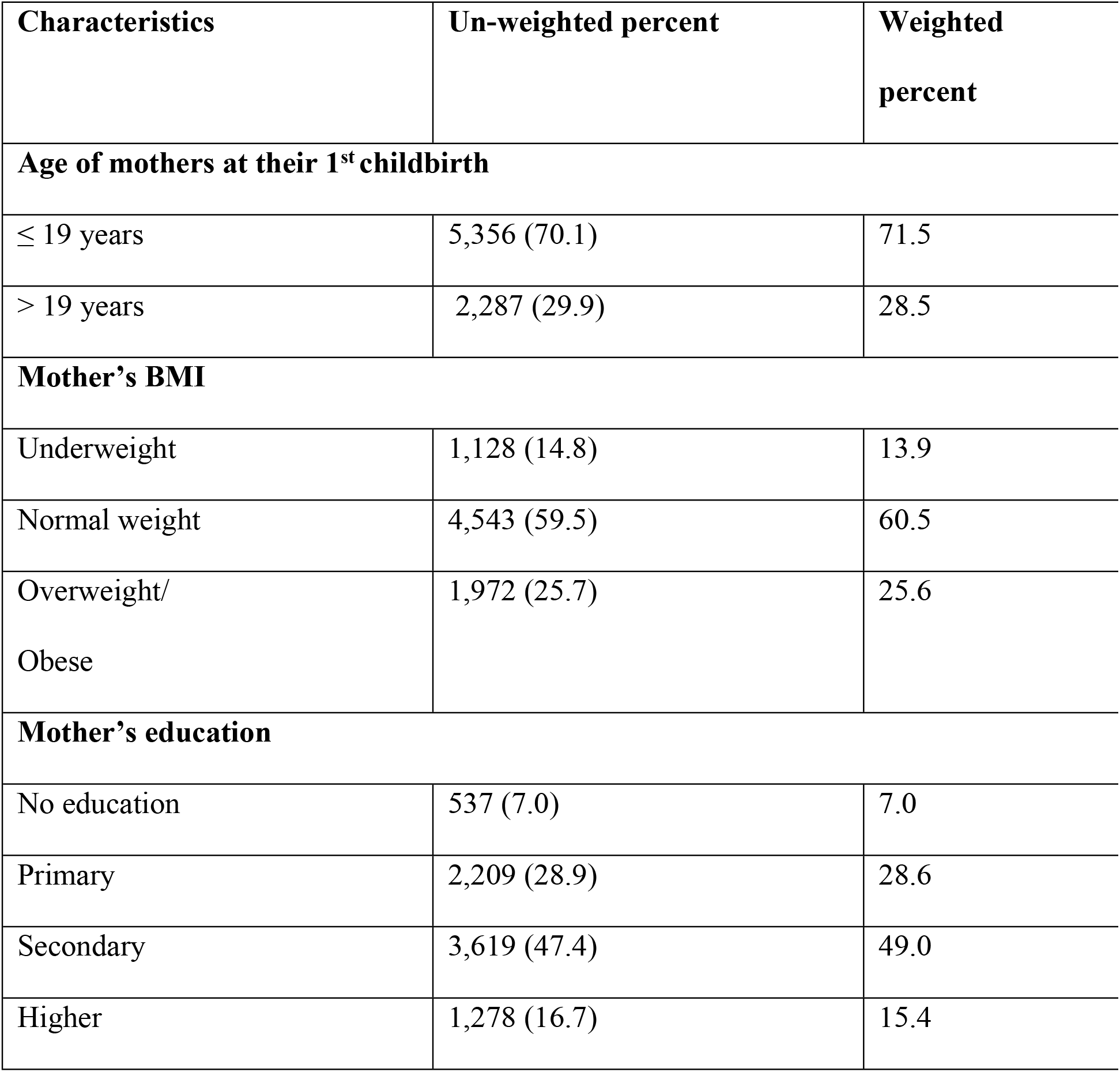
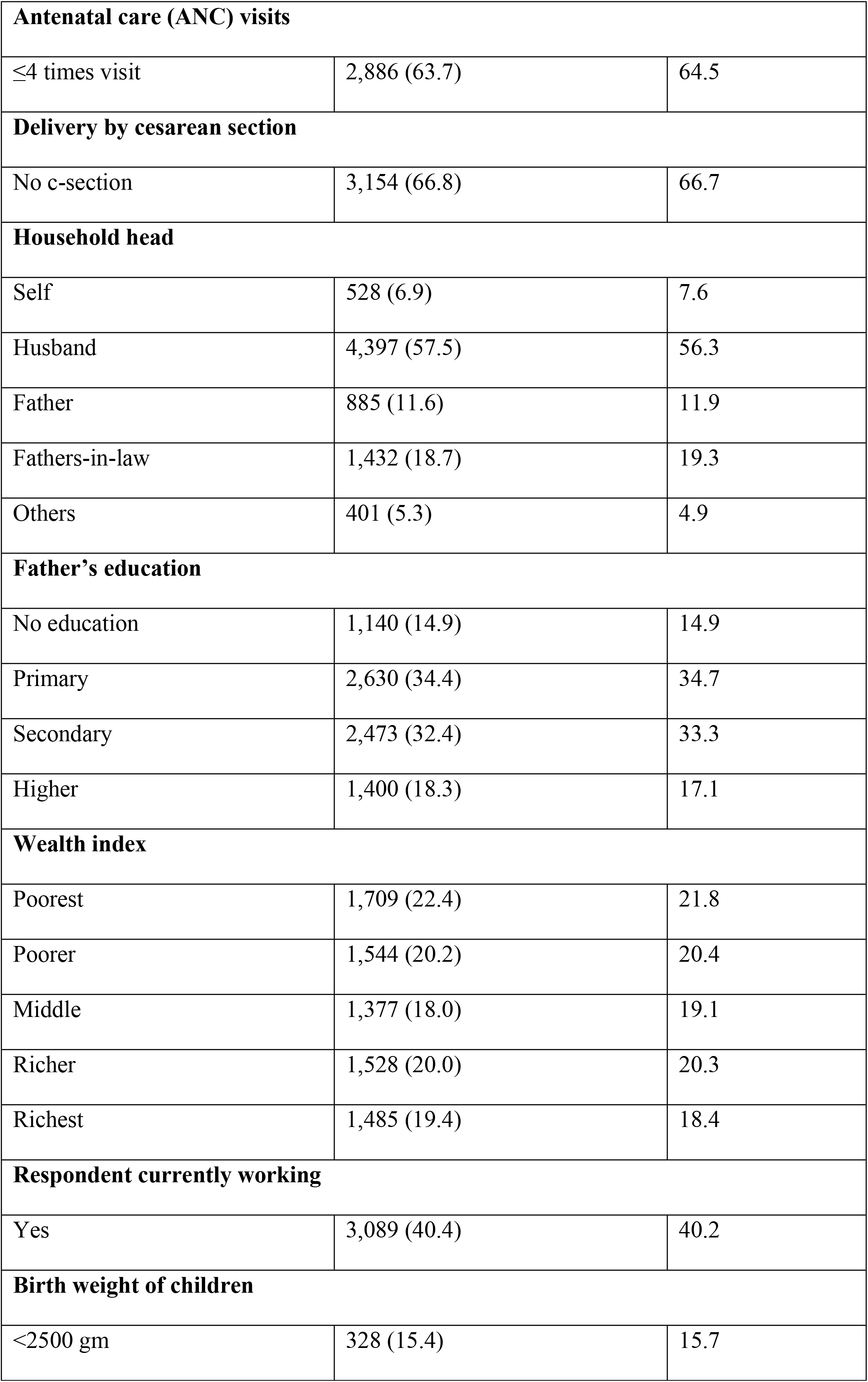
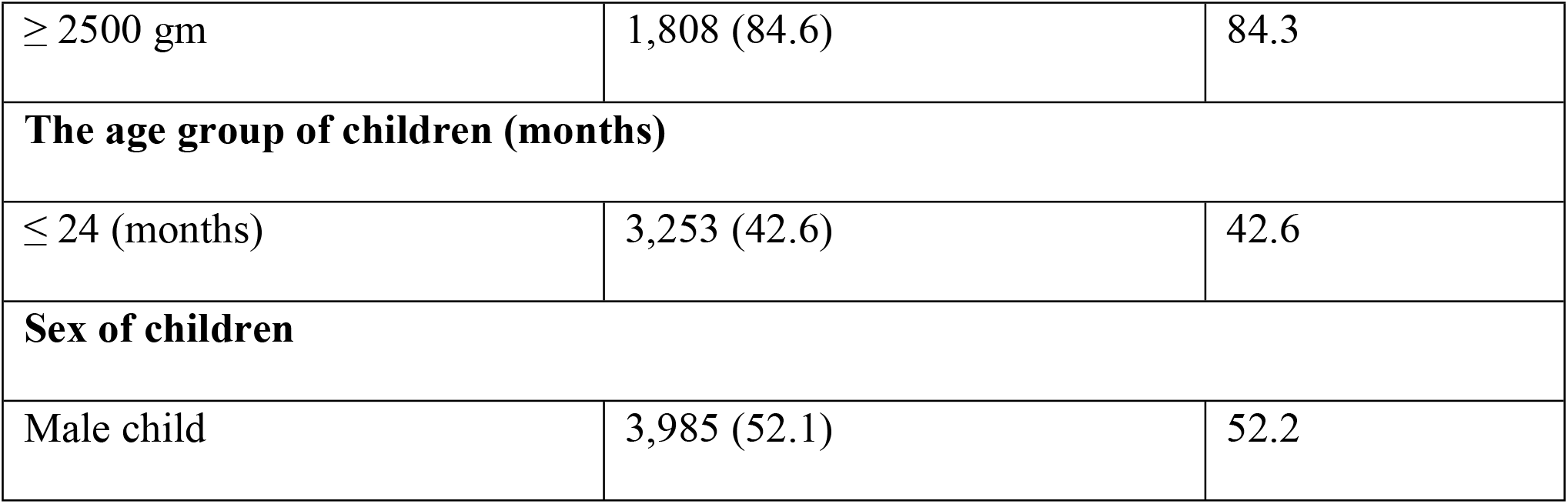
Distribution of background characteristics, presented as number and percentage.

### Nutritional status of children

Stunting, wasting, and underweight were all common, with prevalence rates of 31%, 8%, and 22%, respectively (Figure 2). In a chi-square test, it was found that young mothers’ first-born children had different nutritional statuses from those of adult mothers in terms of underweight (p<0.001), stunted (p<0.001), and wasted (p=0.008). The chi-square test revealed a significant association between child undernutrition and the household head wasted (p=0.017), stunted (p<0.001), underweight (p<0.001), the father’s education wasted (p=0.013), stunted (p<0.001), underweight (p<0.001), and cesarean delivery (p=0.006), wasted (p<0.001), stunted (p<0.001), underweight (p<0.001). Antenatal care visits four times were found to be effective in preventing stunting (p<0.001) and underweight (p<0.001) and but not significant for preventing wasting (p=0.226) (Table 2).

**Table 2:**
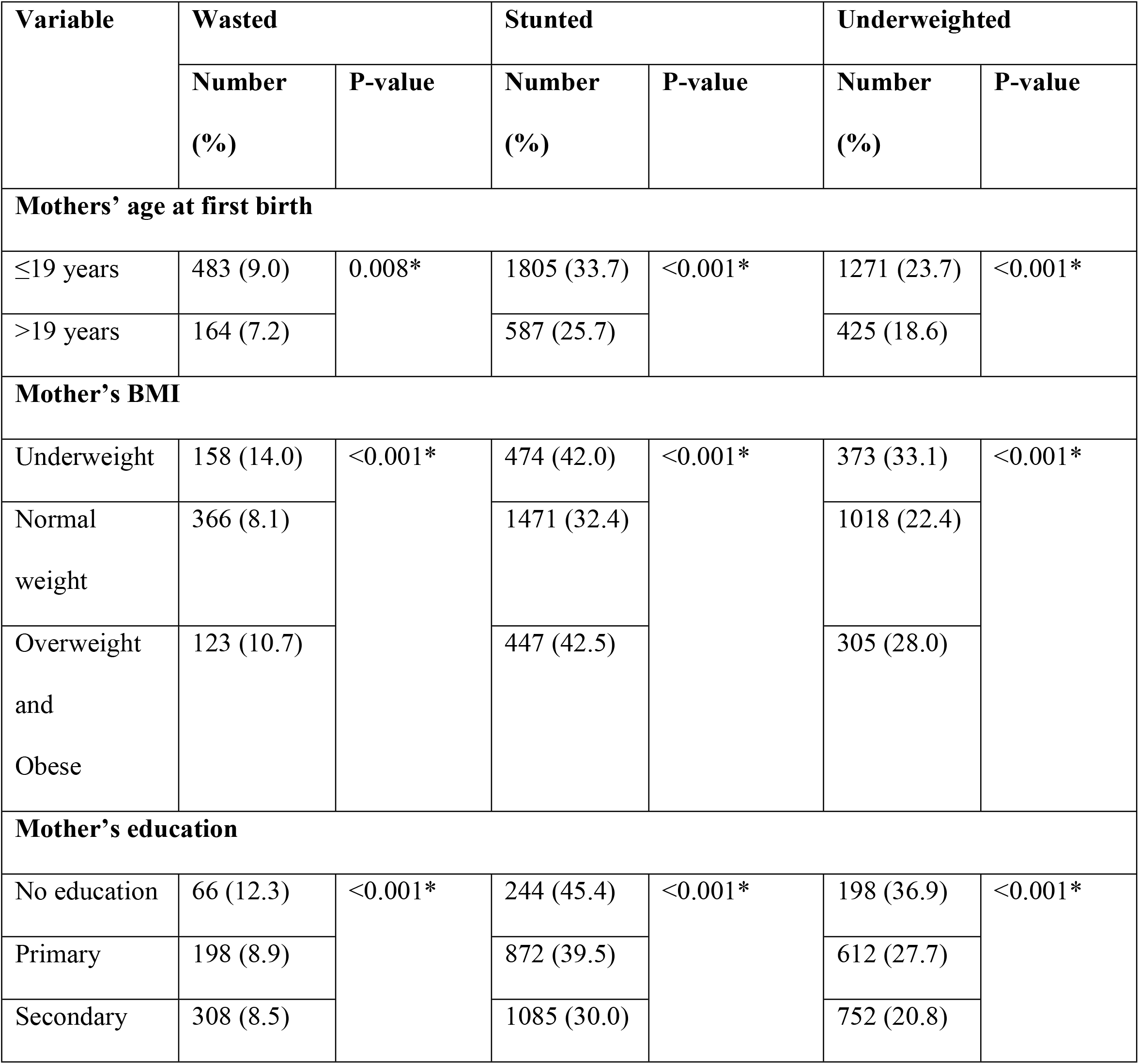
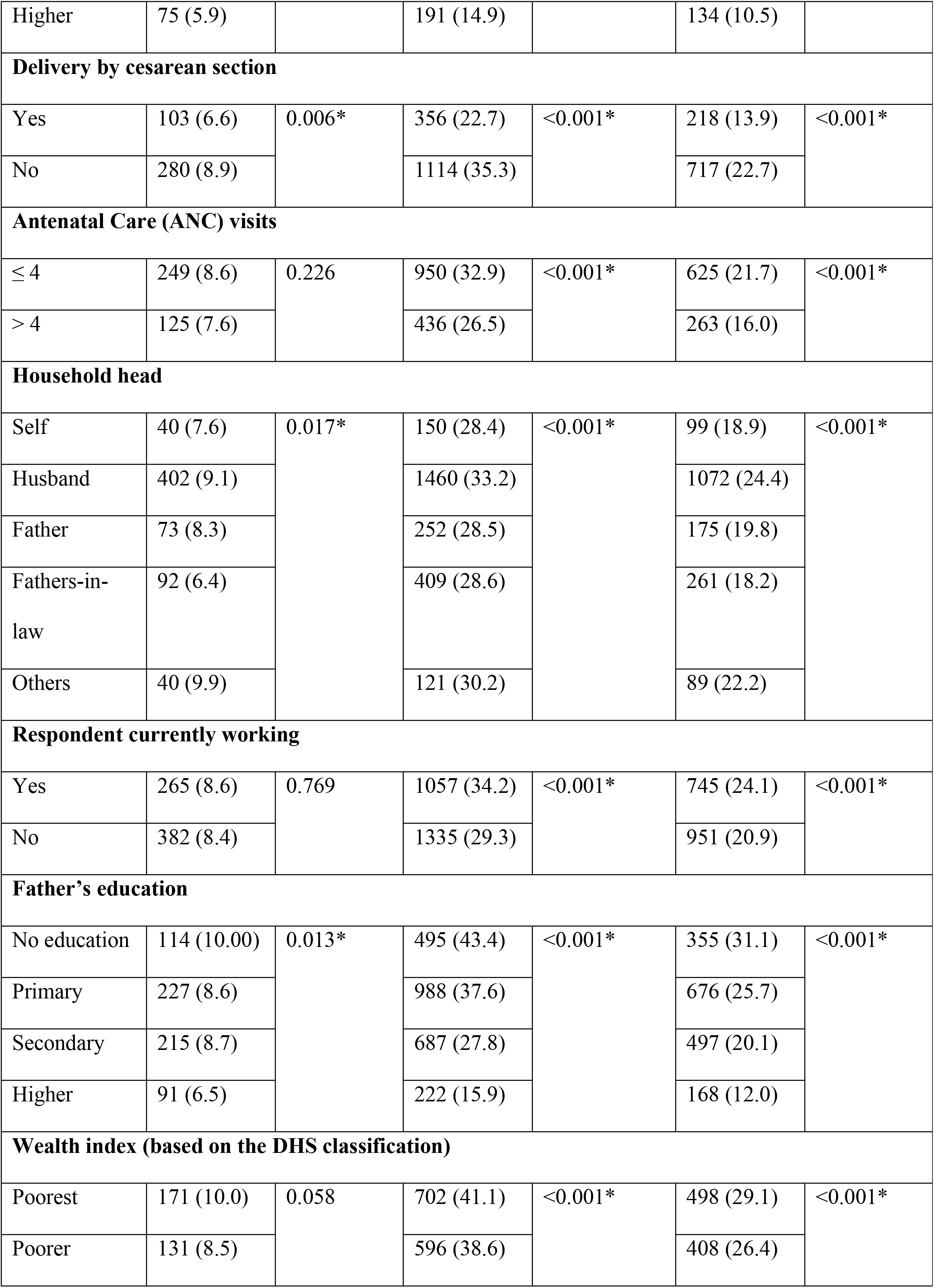
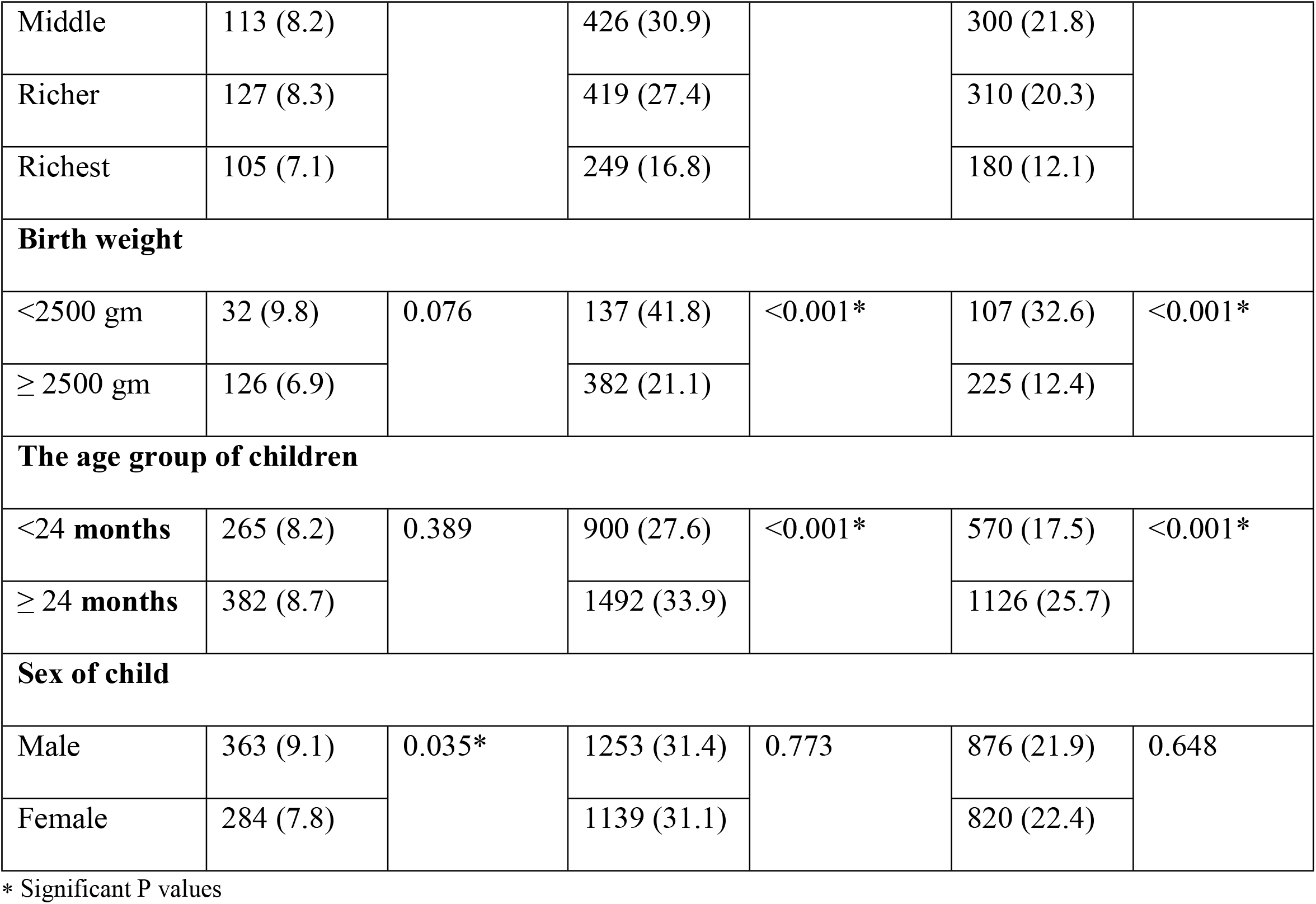
Association of background characteristics and child undernutrition.

| Variable | Wasted |  | Stunted |  | Underweighted |  |
| --- | --- | --- | --- | --- | --- | --- |
|  | Number<br>(%) | P-value | Number<br>(%) | P-value | Number<br>(%) | P-value |
| Mothers' age at first birth |  |  |  |  |  |  |
| ≤19 years | 483 (9.0) | 0.008* | 1805 (33.7) | <0.001* | 1271 (23.7) | <0.001* |
| >19 years | 164 (7.2) |  | 587 (25.7) |  | 425 (18.6) |  |
| Mother's BMI |  |  |  |  |  |  |
| Underweight | 158 (14.0) | <0.001* | 474 (42.0) | <0.001* | 373 (33.1) | <0.001* |
| Normal<br>weight | 366 (8.1) |  | 1471 (32.4) |  | 1018 (22.4) |  |
| Overweight<br>and<br>Obese | 123 (10.7) |  | 447 (42.5) |  | 305 (28.0) |  |
| Mother's education |  |  |  |  |  |  |
| No education | 66 (12.3) | <0.001* | 244 (45.4) | <0.001* | 198 (36.9) | <0.001* |
| Primary | 198 (8.9) |  | 872 (39.5) |  | 612 (27.7) |  |
| Secondary | 308 (8.5) |  | 1085 (30.0) |  | 752 (20.8) |  |
| Higher | 75 (5.9) |  | 191 (14.9) |  | 134 (10.5) |  |
| Delivery by cesarean section |  |  |  |  |  |  |
| Yes | 103 (6.6) | 0.006* | 356 (22.7) | <0.001* | 218 (13.9) | <0.001* |
| No | 280 (8.9) |  | 1114 (35.3) |  | 717 (22.7) |  |
| Antenatal Care (ANC) visits |  |  |  |  |  |  |
| ≤ 4 | 249 (8.6) | 0.226 | 950 (32.9) | <0.001* | 625 (21.7) | <0.001* |
| > 4 | 125 (7.6) |  | 436 (26.5) |  | 263 (16.0) |  |
| Household head |  |  |  |  |  |  |
| Self | 40 (7.6) | 0.017* | 150 (28.4) | <0.001* | 99 (18.9) | <0.001* |
| Husband | 402 (9.1) |  | 1460 (33.2) |  | 1072 (24.4) |  |
| Father | 73 (8.3) |  | 252 (28.5) |  | 175 (19.8) |  |
| Fathers-in-law | 92 (6.4) |  | 409 (28.6) |  | 261 (18.2) |  |
| Others | 40 (9.9) |  | 121 (30.2) |  | 89 (22.2) |  |
| Respondent currently working |  |  |  |  |  |  |
| Yes | 265 (8.6) | 0.769 | 1057 (34.2) | <0.001* | 745 (24.1) | <0.001* |
| No | 382 (8.4) |  | 1335 (29.3) |  | 951 (20.9) |  |
| Father’s education |  |  |  |  |  |  |
| No education | 114 (10.00) | 0.013* | 495 (43.4) | <0.001* | 355 (31.1) | <0.001* |
| Primary | 227 (8.6) |  | 988 (37.6) |  | 676 (25.7) |  |
| Secondary | 215 (8.7) |  | 687 (27.8) |  | 497 (20.1) |  |
| Higher | 91 (6.5) |  | 222 (15.9) |  | 168 (12.0) |  |
| Wealth index (based on the DHS classification) |  |  |  |  |  |  |
| Poorest | 171 (10.0) | 0.058 | 702 (41.1) | <0.001* | 498 (29.1) | <0.001* |
| Poorer | 131 (8.5) |  | 596 (38.6) |  | 408 (26.4) |  |
| Middle | 113 (8.2) |  | 426 (30.9) |  | 300 (21.8) |  |
| Richer | 127 (8.3) |  | 419 (27.4) |  | 310 (20.3) |  |
| Richest | 105 (7.1) |  | 249 (16.8) |  | 180 (12.1) |  |
| Birth weight |  |  |  |  |  |  |
| <2500 gm | 32 (9.8) | 0.076 | 137 (41.8) | <0.001* | 107 (32.6) | <0.001* |
| ≥ 2500 gm | 126 (6.9) |  | 382 (21.1) |  | 225 (12.4) |  |
| The age group of children |  |  |  |  |  |  |
| <24 months | 265 (8.2) | 0.389 | 900 (27.6) | <0.001* | 570 (17.5) | <0.001* |
| ≥ 24 months | 382 (8.7) |  | 1492 (33.9) |  | 1126 (25.7) |  |
| Sex of child |  |  |  |  |  |  |
| Male | 363 (9.1) | 0.035* | 1253 (31.4) | 0.773 | 876 (21.9) | 0.648 |
| Female | 284 (7.8) |  | 1139 (31.1) |  | 820 (22.4) |  |

\* Significant P values

**Fig. 2:**
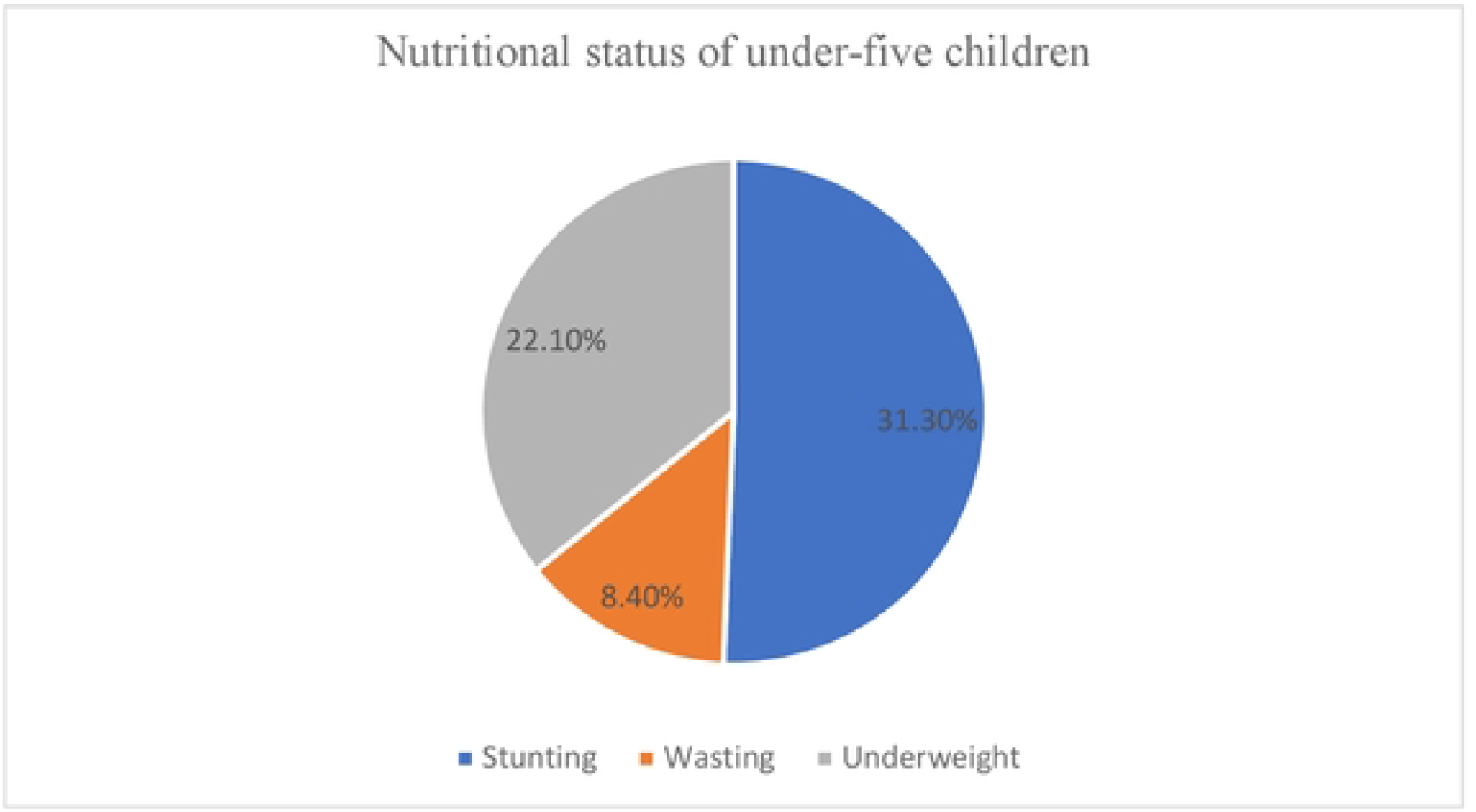
Percentage of undernutrition of under-five children.

### Relationship between mothers’ age during their first delivery and children’s undernutrition

Multiple logistic regression models are used to analyze the data, after adjusting for potential confounders such as household head type, antenatal care (ANC), childbirth weight, and the respondents’ current employment. In the regression model, AOR =1.6, 95% CI 1.09-2.46 were found to be 1.6 times more likely to be wasted, AOR =1.4, 95% CI 1.09-1.78 were found to be 1.4 times more likely to be stunted, and AOR =1.5, 95% CI 1.15-2.07 were found to be 1.5 times more likely to be underweight of children born to mothers aged less than nineteen years compared to mothers aged more than nineteen years (Table 3).

**Table 3:** the relationship between the nutritional state of u-5 children with young and adult mothers.

| Characteristics | Unadjusted model |  | Adjusted model <sup>a</sup> |  |
| --- | --- | --- | --- | --- |
|  | Crude odds ratio<br>(95% CI) | P-value | Adjusted odds<br>ratio (95% CI) | P-value |
| **Wasting** |  |  |  |  |
| >19 years | Reference |  | Reference |  |
| ≤19 years | 1.3 (1.03,1.61) | 0.024* | 1.6 (1.02, 2.46) | 0.039* |
| **Stunting** |  |  |  |  |
| >19 years | Reference |  | Reference |  |
| ≤19 years | 1.4 (1.23,1.59) | <0.001* | 1.4 (1.09,1.78) | 0.008* |
| **Underweight** |  |  |  |  |
| >19 years | Reference |  | Reference |  |
| ≤19 years | 1.3 (1.14,1.52) | <0.001* | 1.5 (1.15,2.07) | 0.004* |

<sup>a</sup> Model adjusted for ANC visit , child birth weight, respondent currently working and type of household head
\* Significant P values

## Discussion

This study found that 71.5% of women who gave birth for the first time were below the 19 years. The prevalence of maternal age at first birth in Bangladesh aligns with the previous study in Bangladesh (29). In Bangladesh, marriage at a young age is forbidden. However, it still exists due to religious and societal factors, peer pressure, perceptions of girls’ gender roles, upholding family honor and virginity, dowry, and concern about safety and security (31-33). According to Bangladeshi civil law, girls must be 18 years to get married. In 2017; the Bangladeshi parliament approved early marriage in certain situations (34). As a result, many young girls married before their legal marriage age limit. In Bangladesh, 33% of young mother marriages before the age of 15, and 66% of young mother marriages before age the of 18 years (35). Sixty-eight percent of girls had a child before twenty years of age (age 15-19 years) (36). Because of this, many women have children before they were 19 years old. That has detrimental effects on raising and caring for children. In USA, a study shows that experiences for young mothers on childbearing have some effects on their minds, relationships, and emotions (37).

This research revealed that higher risk of stunting, wasting, and underweight among under-five children whose mothers were age of below 19 years. According to this study, having the first child at the age before 19 years puts at an elevated risk of child undernutrition, including stunting and wasting. Children of adolescent mothers are more likely to be undernourished such as shorter and underweight compare to the adult mothers (38). Young pregnant mothers are associated with undernutrition in children through poor maternal nutrition, less education, less access to health services, poor supplementary feeding practices, and substandard housing situations (12). A prospective study found that young maternal age (≤ 19 years) is associated with the stunting of two years aged children (27).

The overall prevalence of stunting, wasting, and underweight in under-five children of young mothers is 33.7%, 9.0%, and 23.7%, respectively. Another study conducted in Bangladesh on maternal, newborn, and child health (MNCH) programs area of BRAC (a non-governmental organization) found the prevalence of stunting 15.9%, wasting 14.7%, and underweight 22.4%, respectively of the children of mother age less than nineteen years old (39). In Ghana, under-five children of young maternal age (15-19 years) are more likely to be stunted (39.0%) and wasted (8.0%) (40).

Current study found that children of young mothers (age below 19 years) more likely to be undernutrition than children with older mothers. These children were at least 1.4 times more likely than children of adult mothers to be underweight, stunted, and wasting. In India, a study found that young pregnancies were linked to an increased risk of undernutrition. Children of young mothers were five percentage points more likely to be stunted than children of adult mothers, and they were also shorter for their age (41).

In this study, it is found that mother who gave their first birth at an age below 19 years is more accountable for the undernutrition of children. A review found that mothers who gave birth at the age of before 19 years received routine medical care and immunizations for children, frequently overfed or underfed their children, fewer mothers breastfed their children and they introduce solid foods in the first or second months of the infants (18). This occurs among mothers below nineteen years, who may not be able to nurture appropriately due to the lack of their maturity. Many adolescent mothers had lack of knowledge to take responsibility for maternity and infant care including prenatal care, and breastfeeding (42).

Maternal age at first birth of under 19 years who get pregnant too young and leave school may feel personally unhappy if they don’t have a steady source of income. Pregnant girls are frequently forced or under pressure to leave school, which might affect their chances for higher education and jobs (43). Due to the early pregnancy, they might give their kids less care, breastfeeding, and nurturing than mothers who are adults, which might translate to less care, nursing, and nurturing all around (42). Adolescent girls have been negatively impacted on their couples, families, schools, and society at large by early motherhood (42). Due to malnutrition and other growth deficiencies, this is likely to have an impact on the physical and cognitive development of their children (44).

### Strengths and Weakness

This study analyzes nationally representative demographic and health surveys data to examine the relationship between maternal age below 19 years and undernutrition of under-five children. BDHS-17/18 data helps to generalize the relationship between the maternal age first birth and undernutrition of under-five children at the national level.

We performed a cross-sectional analysis that is not appropriate for studying causal relationships but it shows that the association between the maternal age at first birth and undernutrition of under-five children. In contrast, data were collected on mothers who have given birth at the age below nineteen years using the recall procedure so that it may have some remembrance issues.

## Data Availability

https://dhsprogram.com/data/dataset_admin/index.cfm

## Conclusion and recommendations

These research results provide policymakers with vital information to implement the programs focusing on child undernutrition and younger maternal age at first birth ≤ 19 years. Moreover, young mother need to engage with the inclusive education to reduce the challenges of child health and nutrition. A longitudinal investigation is necessary to determine the causal connection between the maternal age at first birth and the undernutrition of under-five children.

Additionally, young mothers need special program during pregnancy and the postpartum period. Moreover, a qualitative study might be helpful to understand the sociocultural factors that related with young maternal age and child undernutrition.

## Acknowledgments

We appreciate the permission from the Bangladesh National Institute of Population Research and Training (NIPORT) and MEASURE DHS to utilize the BDHS dataset. Moreover, we specially thanks to our core donor Government of Bangladesh, and Canada for their support.

## Availability of data

Available at: https://dhsprogram.com/data/dataset_admin/index.cfm

## Notes

### Competing Interest Statement

The authors have declared no competing interest.

### Funding Statement

The author(s) received no specific funding for this work.

### Author Declarations

DHS data are publicly available upon justifiable request and are in the public domain. We did not need further ethical approval because the study protocol was accepted by both the Bangladeshi Ethics Commission and ICF International. No animal data are used.

## References

1. Maleta K. Undernutrition. Malawi Medical Journal. 2006;18(4).

2. Nutrition [Internet]. 2022 [cited April 2, 2022]. Available from: https://www.who.int/health-topics/nutrition.

3. Malnutrition [Internet]. 2021 [cited April 2, 2022]. Available from: https://www.who.int/news-room/fact-sheets/detail/malnutrition.

4. WHO. Director-General’s opening remarks at the second informal member state briefing on UN Food Systems Summit. April 21, 2021 [Available from: https://www.who.int/director-general/speeches/detail/director-general-s-opening-remarks-at-the-second-informal-member-state-briefing-on-un-food-systems-summit.

5. Service WHE. ASIA HUNGER FACTS: What is the extent of hunger in Asia? [Available from: https://www.worldhunger.org/asia-hunger-facts/.

6. South-eastern Asia; The burden of malnutrition at a glance [Internet]. 2021 [cited April 2, 2022]. Available from: https://globalnutritionreport.org/resources/nutrition-profiles/asia/south-eastern-asia/.

7. National Institute of Population Research and Training - NIPORT MoHaFW, and ICF. 2020. Bangladesh Demographic and Health Survey 2017-18. 2020.

8. Bangladesh; The burden of malnutrition at a glance [Internet]. 2022. Available from: https://globalnutritionreport.org/resources/nutrition-profiles/asia/southern-asia/bangladesh/.

9. Mark Myatt TK, Simon Schoenbuchner, Silke Pietzsch, Carmel Dolan, Natasha Lelijveld, and André Briend. Children who are both wasted and stunted are also underweight and have a high risk of death: a descriptive epidemiology of multiple anthropometric deficits using data from 51 countries. Archives of Public Health. 2018;76;28.

10. Cesar G Victora LA, Caroline Fall, Pedro C Hallal, Reynaldo Martorell, Linda Richter, Harshpal Singh Sachdev. Maternal and child undernutrition: consequences for adult health and human capital. 2008.

11. Vinicius J. B. Martins Tmmtf, Luciane P. Grillo, Maria Do Carmo P. Franco, Paula A. Martins, Ana Paula G. Clemente, Carla D. L. Santos, Maria de Fatima A. Vieira, Ana Lydia Sawaya. Long-Lasting Effects of Undernutrition. IJERPH. 2011;8.

12. Phuong Hong Nguyen SS, Sumanta Neupane, Lan Mai Tran, Purnima Menon. Social, biological, and programmatic factors linking adolescent pregnancy and early childhood undernutrition: a path analysis of India’s 2016 National Family and Health Survey. The Lancet Child & adolescent health. 2019.

13. Organization WH. Adolescent Pregnancy Issues in Adolescent Health and Development. 2004.

14. Adolescent pregnancy [Internet]. 2023 [cited July 12, 2023]. Available from: https://www.who.int/news-room/fact-sheets/detail/adolescent-pregnancy.

15. Adolescents in South Asia [Internet]. 2021. Available from: https://www.unicef.org/rosa/what-we-do/adolescents.

16. Icrw WB, editor Economic Impacts of Child Marriage: Global Synthesis Report 2017.

17. Kassebaum N KH, Zoeckler L, Olsen HE, Thomas K, Pinho C, Bhutta ZA, Dandona L, Ferrari A, Ghiwot TT, Hay SI. Child and Adolescent Health From 1990 to 2015 Findings From the Global Burden of Diseases, Injuries, and Risk Factors 2015 Study. 2017.

18. Hechtman L. Teenage mothers and their children: risks and problems: a review. 1989.

19. HELEN M. Dijplessis RB, TONI Richards. Adolescent Pregnancy: Understanding the Age and Race on Outcomes 1997.

20. Soo Hyun Yu JM, Jennifer Crum, Claudia Cappa, David R. Hotchkiss. Differential effects of young maternal age on child growth. 2016.

21. Organization WH. Adolescent pregnancy situation in South-East Asia Region. 2014.

22. Organization WH. Preventing early pregnancy and poor reproductive outcomes among adolescents in developing countries: what the evidence says. 2011.

23. Organization WH. Women and health : today’s evidence tomorrow’s agenda. 2009.

24. Heather L. Sipsma UM, Anna Divney, Derrick Gordon, Elizabeth Gabzdyl, Trace Kershaw. Breastfeeding behavior among adolescents: Initiation, duration, and exclusivity. 2013.

25. T K LeGrand CSM. Teenage Pregnancy and Child Health in the Urban Sahel. 1993.

26. Srinivas Goli AR, Deepti Singh. The Effect of Early Marriages and Early Childbearing on Women’s Nutritional Status in India. 2015.

27. Fall CH SH, Osmond C, Restrepo-Mendez MC, Victora C, Martorell R, Stein AD, Sinha S, Tandon N, Adair L, Bas I, Norris S, Richter LM; COHORTS investigators. Association between maternal age at childbirth and child and adult outcomes in the offspring: a prospective study in five low-income and middle-income countries (COHORTS collaboration). Lancet Glob Health. 2015.

28. Tahmeed Ahmed MH, Kazi Istiaque Sanin, . Global Burden of Maternal and Child Undernutrition and Micronutrient Deficiencies Annals of nutrition & metabolism. 2013;61:8–17.

29. Phuong Hong Nguyen SS, Long Quynh Khuong, Priyanjana Pramanik, Akhter Ahmed, Sabina Faiz Rashid, Kaosar Afsana, and Purnima Menon. Adolescent birth and child undernutrition: an analysis of demographic and health surveys in Bangladesh, 1996–2017. Annals of the New York Academy of Sciences. 2021.

30. WHO. The WHO Child Growth Standards. 2010.

31. Chowdhury FD. The socio-cultural context of child marriage in a Bangladeshi village. International Journal of Social Welfare. 2004;13(3):244–253.

32. Elizabeth G Henry NBL, Ashraful Alam, Nabeel Ashraf Ali, Emma K Williams, Syed Moshfiqur Rahman, Salahuddin Ahmed, Shams El Arifeen, Abdullah H Baqui, Peter J Winch. Sociocultural factors perpetuating the practices of early marriage and childbirth in Sylhet District, Bangladesh. 2015.

33. Rahman M. Determinates of early marriage in Bangladesh: An evidence of the nationally representative survey. International Journal of Sociology and Anthropology. 2017.

34. WATCH HW. Bangladesh: Legalizing Child Marriage Threatens Girls’ Safety Contain Harm with Strict Regulations. 2017 [Available from: https://www.hrw.org/news/2017/03/02/bangladesh-legalizing-child-marriage-threatens-girls-safety.

35. Matera B. New Bangladesh marriage law is blow to children’s and women’s rights. 2014.

36. Mohammad Mainul Islam MKI, Mohammad Sazzad Hasan, Mohammad Bellal Hossain. Adolescent motherhood in Bangladesh:Trends and determinants. 2017.

37. Mollborn S. Teenage Mothers Today: What We Know and How It Matters. 2016.

38. Phuong Hong Nguyen SS, Sumanta Neupane, Lan Mai Tran, Purnima Menon. Social, biological, and programmatic factors linking adolescent pregnancy and early childhood undernutrition: a path analysis of India’s 2016 National Family and Health Survey. The Lancet Child & Adolescent Health. 2019;3(7):P463–73.

39. Phuong Hong Nguyen TS, Lan Mai Tran, Kaosar Afsana, Zeba Mahmud, Bachera Aktar, Raisul Haque, Purnima Menon. The nutrition and health risks faced by pregnant adolescents: Insights from a cross-sectional study in Bangladesh. PLoS ONE. 2017.

40. Anthony Wemakor HG, Thomas Azongo, Helene Garti, Ambrose Atosona. Young maternal age is a risk factor for child undernutrition in Tamale Metropolis, Ghana. BMC research notes. 2018.

41. Bhan N. Preventing teenage pregnancy in India to end the cycle of undernutrition. The Lancet. 2019;3.

42. Massoumeh Mangeli MR, Mohammad Ali Cheraghi, and Batool Tirgari,. Exploring the Challenges of Adolescent Mothers From Their Life Experiences in the Transition to Motherhood: A Qualitative Study. J Family Reprod Health. 2017.

43. Early childbearing [Internet]. 2022. Available from: https://data.unicef.org/topic/child-health/adolescent-health/.

44. THE CHANGING FACE OF MALNUTRITION: THE STATE OF THE WORLD’S CHILDREN 2019 [Internet]. [cited 31 July 2023]. Available from: https://features.unicef.org/state-of-the-worlds-children-2019-nutrition/.

